# Interaction of Heavy Drinking Patterns and Depression Severity predicts Efficacy of Quetiapine Fumarate XR in lowering Alcohol Intake in Alcohol Use Disorder Patients

**DOI:** 10.1101/2020.07.01.20144311

**Authors:** Vatsalya Vatsalya, Maiying Kong, Luis M. Marsano, Zimple Kurlawala, Kan V. Chandras, Melanie L. Schwandt, Vijay A. Ramchandani, Craig J. McClain

**Author notes:** **Corresponding Author Address:** Vatsalya Vatsalya MD, MS, PgD, MSc, Department of Medicine, University of Louisville, 505 S. Hancock St., CTR Room 514A, Louisville KY 40202.

## Abstract

**Background:** Shared etiological pathways of dopamine and serotonin neurotransmission play a central role in heavy alcohol intake and exacerbation in the symptoms of depression.

We investigated the role of depression ratings and patterns of heavy drinking on the treatment efficacy of Quetiapine fumarate XR in lowering alcohol intake in alcohol use disorder (AUD) patients.

**Methods:** One hundred and eight male and female heavy drinking AUD patients in the age range of 18–64 yrs. received 12 weeks of active treatment. Participants were grouped by the severity grading of depression using Montgomery-Asberg Depression Rating Scale (MADRS) (clinically relevant≥8 [CR], clinically non-relevant≤7 [CNR]) at baseline. Drinking history and depression ratings were assessed at the patients visits.

**Results:** Heavy drinking days (HDD) and total drinks (TD) were significantly fewer in CR patients at the treatment end. A true positive response in AUROC analysis supported the lowering of TD in CR patients. The number of drinking days (NDD) and average drinks per drinking day (AvgD) were lower in the CNR patients at treatment-end. Significant associations with increasing effect sizes were observed for all the heavy drinking measures (HDD, TD, NDD and AvgD) and MADRS scores by the end of the treatment course.

**Conclusions:** Baseline elevated depressive symptoms could likely predict the course of heavy alcohol drinking during the treatment, and efficacy outcome of a treatment. AUD patients with baseline clinically significant depression had a progressive lowering in heavy drinking markers significantly corresponding to the lowering of depression symptoms by the end of treatment with Quetiapine fumarate XR.

**ClinicalTrials.gov:** NCT#0049862 (https://clinicaltrials.gov/ct2/show/NCT00498628?term=litten&draw=2&rank=3)

## Introduction

Alcohol use disorder (AUD) is an important mental and medical health concern in the United States. Heavy prolonged alcohol intake can cause organ injury, including neurodegenerative and adverse neurocognitive effects. ^1^ Currently, the FDA approved medications for treatment of alcohol use disorder (AUD), i.e. disulfiram, acamprosate, and oral and injectable naltrexone ^2,3^, have not been proven to be efficacious for all patients. Around 30% of the patients with bipolar depression and schizophrenia conditions report higher susceptibility to heavy alcohol drinking. ^4,5^ Heavy alcohol intake and depression have been shown to have close comorbid susceptibility thus there is a high likelihood finding depression symptoms in alcohol dependents. ^6^

Some studies have shown this propensity with the involvement of dopamine D2 and serotonin 5-HT2 pathways in such mental comorbid conditions along with heavy alcohol intake ^7,8^. This propensity of alcohol and mental condition has been mechanistically explained using the same pathways both in human and animal studies. ^9-11^ Alcohol intake in excessive amounts could also adversely impact the prognosis in these conditions. ^12,13^ Among the clinical trials conducted to date, Quetiapine has been shown to have positive effects on alcohol consumption--increased days of abstinence from alcohol during the 2–7 months of treatment ^14^; increased abstinent days and improvement in depression, anxiety, and insomnia ^15^; and increased abstinent days and fewer hospitalizations in alcohol-dependent patients with disturbed sleep. ^16^ Such mental conditions are difficult to manage and have shown broad co-morbidity. ^17^

Heavy alcohol drinking has been well characterized by the recent drinking history using Timeline Followback (TLFB) for past 90 days in AUD patients in recent research on AUD. ^18,19^ These markers have also been tested on AUD patient cohorts who also exhibited other form of addiction such as cocaine, pathophysiological or behavioral symptomology including depression. ^20-22^ Heavy drinking markers derived from TLFB do not only characterize the drinking behavior, consequences, and pathological course but are also highly useful as therapeutic targets of treatment efficacy. ^23,24^ Thus, in this study, we used TLFB as our primary assessment of endpoints for treatment efficacy

A pharmacotherapeutic drug that could reduce alcohol consumption and adequately manage complications of chronic alcohol consumption would be a desirable treatment alternate for reducing alcohol intake. A pilot study had shown lowering of alcohol consumption with Quetiapine treatment in heavy drinkers. ^25^ The preliminary results described above suggest that Quetiapine may have potential in treatment for alcohol dependence, especially among heavy drinkers. Quetiapine is an FDA approved treatment for depression, bipolar disorder, and schizophrenia. Thus, we postulated that Quetiapine may be a useful intervention to reduce heavy alcohol drinking in alcohol dependents who also exhibit symptoms of depression following previous literature. ^6,26^

This study was conducted as a secondary analysis of a larger clinical trial on quetiapine efficacy for reducing alcohol consumption. ^27^ Primary aim of this study is to determine if baseline and longitudinally reported depression ratings and corresponding heavy drinking patterns could predict efficacy of Quetiapine XR in reducing heavy alcohol drinking to a moderate level. Our other aim was to identify the specific markers/patterns of heavy drinking that show affinity with the symptoms of depression in AUD patients.

## Methods

### Study Participants

This study is one of the investigational arms of a larger protocol (ClinicalTrials.gov: NCT#00498628) that was supported by National Institute on Alcohol Abuse and Alcoholism (NIAAA). This investigation was a double-blind placebo-controlled parallel group design with two treatment groups: (1) Quetiapine XR as an active drug, and (2) placebo as control that was approved by the institutional review board of all the participating sites. One hundred and seventy-nine men and 45 women were randomized in this larger study after the completion of consenting, and 218 started the treatment. ^27^ One hundred and eight patients who received Quetiapine fumarate XR among the total randomized were included in our study.

Inclusion criteria included diagnosis with alcohol dependence (using Diagnostic and Statistical Manual of Mental Disorders, Fourth Edition) and age between 18–64 years. Other inclusion criteria were 10 or more drinks per day for men and 8 or more drinks per day for women for at least 40% of the last 60 days of the 90-day drinking assessment (Time-line Follow-back, TLFB90). Having a 0.00 breath alcohol level at the time of consenting was also a requirement. Major exclusions were: other psychoactive drug dependence within the last year, positive urine screen for drugs, participation in other pharmacological/behavioral study within the last three months, lifetime diagnosis of major depression or eating disorder, use of antidepressants (last 30 days) and antipsychotics (last 14 days) before randomization, other significant medical conditions such as panic disorder with or without agoraphobia, schizophrenia, bipolar disorder, or other psychosis, or a past-year diagnosis of major depression or eating disorder, and Clinical Institute Withdrawal Assessment of Alcohol score ≥10.

### Procedures and Assessments

The active drug, Quetiapine XR (Seroquel XR® AstraZeneca, Wilmington DE) was provided to the participants for three months in 50- and 200-mg tablets with identical matching non-active pills for the placebo group. ^27^ Blood chemistry; clinical and subjective assessments from baseline (0W, start of the trial treatment); end of four-week (4W, post 3-weeks of dose titration or dose escalation); end of eight weeks (8W, mid of maintenance phase of dosing); and end of week 12 (12W, end of maintenance phase of dosing when dose tapering phase commenced) were evaluated. Dose was titrated in the first three weeks up to a target dose of 400 mg/d, which was maintained from the start of week 4 until the end of week 12. All individuals received medical management (MM) that included assessment of medication side effects, participant education and advice on drinking. ^28^

### Data Collection, Statistical Paradigm and Analysis

Individual demographics—age (years), sex (male or female), weight (lbs.), and drinking history (TLFB90) were collected at the time of screening evaluation and were included in this study to estimate their role as pre-existing conditions and factors in the Quetiapine XR pharmacodynamics. Recent TLFB90 measures ^29^ developed from the raw data included Total Drinks (TD90), Drinking Days past 90 Days (NDD90), Average Drinks per Drinking Day in past 90 Days (AvgDPD90), and Heavy Drinking Days (defined as five or more drinks per day for a man and four or more drinks per day for a woman) in the past 90 Days (HDD90). Additionally, we analyzed drinking history at each timepoint : -2–0W (between screening and baseline for two weeks); 4W (four weeks, from baseline to the end of week 4); 8W (two weeks, from the beginning of the 7^th^ week to the end of 8^th^ week); and 12W (two weeks, starting from the beginning of the 11^th^ week to the end of 12^th^ week). We also used the TLFB questionnaire during the treatment period they have been termed as following: Total Drinks (TD), Drinking Days (NDD), Average Drinks per Drinking Day (AvgD), and Heavy Drinking Days (defined as five or more drinks per day for a man and four or more drinks per day for a woman, HDD).

Demographic and drinking history assessment were performed using univariate analysis in the Quetiapine treated and placebo group patients. The MADRS scale was included in this study as the primary tool for grouping the AD patients as having clinically not relevant (CNR) or clinically relevant (CR) depression, based on the MADRS score reported at baseline. MADRS was selected as an instrument for assessment of depression symptoms since it has only one item pertaining to sleep disturbance, due to which clinical investigations and regulatory authorities favor the MADRS to discriminate any prominent nonspecific sedative effects. ^30^ The scale was constructed to be sensitive to changes in treatment effects. Its capacity to differentiate between responders and non-responders to antidepressant treatment has been shown to be comparable to the Hamilton Rating Scale for Depression ^31^, another established measure of depressive symptomatology, but the MADRS has greater sensitivity to change during the course. It has exhibited high inter-rater reliability and appears to be oriented more towards psychic as opposed to somatic aspects of depression. ^32^

Participants were grouped by the severity grading of depression using Montgomery-Asberg Depression Rating Scale (MADRS) (clinically relevant≥8 [CR], clinically non-relevant≤7 [CNR]) at baseline in both the active and placebo groups. Since there were some patients who could have taken antidepressants more than 30 days back (not an exclusion criteria), we also included the between the scale steps’ score for each question (1,3,5) to include any subtle elevations (thus ≤7) in place of defined scale steps (0,2,4,6) for patients ^33^. This assessment was used in various between-group univariate, and repeated ANOVA analyses. MADRS scores were used as covariates in the between group (and by time-course analyses, as applicable) for multiple comparisons. Multiple comparisons were conducted to estimate the increase in effect sizes of the outcome measures (post-treatment drinking markers) only. Association analyses were conducted using linear and multivariable regression models. Data analysis platforms used in this study were SPSS 26.0 version (IBM, Chicago IL), MS Office 365 (MS Corp. Redmond WA) and GraphPad prism 7 (GraphPad Software, Inc., La Jolla CA). Figures were created using MS Office PowerPoint 2016 (MS Corp. Redmond WA) and GraphPad prism 7 (GraphPad Software, Inc., La Jolla CA), and converted to TIFF file. Statistical significance was set at p <0.05. Data are presented as Mean ± Standard Deviation (M±SD).

## Results

### Patient Characterization and Drinking Profile

Demographic measures were comparably similar in all the subgroups differentiated by treatment, and sex (Table 1). As anticipated, males weighed more and numbered more than females in each arm. There was no significant main effect of any of the demographic or drinking history markers in these analyses. There were no statistical differences at baseline in the heavy drinking markers between the AD patients with clinically relevant (CR) depression and those without (CNR) depression.

**Table 1:** Baseline demographic and drinking history markers in alcohol dependent patients by MADRS group and sex.

| Treatment and Measures | Quetiapine |  |  |  | Placebo |  |  |  |
| --- | --- | --- | --- | --- | --- | --- | --- | --- |
|  | CNR |  | CR MADRS |  | CNR |  | CR MADRS |  |
|  | Males (63) | Females (14) | Males (27) | Females (4) | Males (56) | Females (15) | Males (32) | Females (12) |
| Age(yrs.) | 44.4±10.0 | 49.9±8.5 | 46.0±8.1 | 41.8±4.3 | 46.7±9.7 | 46.8±5.7 | 42.3±10.4 | 46.4±11.4 |
| Weight(lb.) | 194.9±40.8 | 153.2±27.6 | 195.1±35.0 | 174.3±35.4 | 201.1±45.5 | 162.0±31.8 | 198.0±47.1 | 163.0±38.5 |
| Drinking History |  |  |  |  |  |  |  |  |
| TD90 | 1268.9±503.5 | 1030.0±507.8 | 1370.0±587.3 | 1064.8±309.1 | 1206.2±461.9 | 949.3±358.4 | 1242.8±479.5 | 955.2±255.0 |
| AvgD90 | 14.1±5.6 | 11.4±5.6 | 15.2±6.5 | 11.8±3.4 | 13.4±5.1 | 10.6±4.0 | 13.8±5.4 | 10.6±2.8 |
| HDD90 | 63.1±22.7 | 72.1±13.1 | 67.7±24.7 | 76.0±16.2 | 68.6±21.5 | 66.9±24.5 | 70.5±19.4 | 73.1±22.3 |
| NDD90 | 80.5±15.1 | 81.3±13.7 | 79.1±13.6 | 83.8±6.5 | 79.6±14.4 | 83.5±7.7 | 79.5±14.1 | 81.9±15.0 |
| Depression Assessment |  |  |  |  |  |  |  |  |
| MADRS | 2.71±2.4 | 3.14±2.1 | 13.48±4.0 | 16.0±5.7 | 2.7±2.3 | 2.40±2.4 | 13.22±4.6 | 12.58±5.2 |

TD90: Total drinks past 90 days; AvgD90: Average drinks per drinking day past 90 days; HDD90: heavy drinking days past 90 days; NDD90: number of drinking days past last 90 days.

### Changes in heavy drinking patterns in Quetiapine treated AD patients

There was no difference in any of the drinking markers recorded at baseline (−2– 0 W) between the CR and CNR groups (Fig. 1a, 2a, 3a, 4a). We found a time-dependent response in the drinking patterns between the CR group compared to the CNR group across all the timepoints (Fig. 1-4).

**Figure 1:**
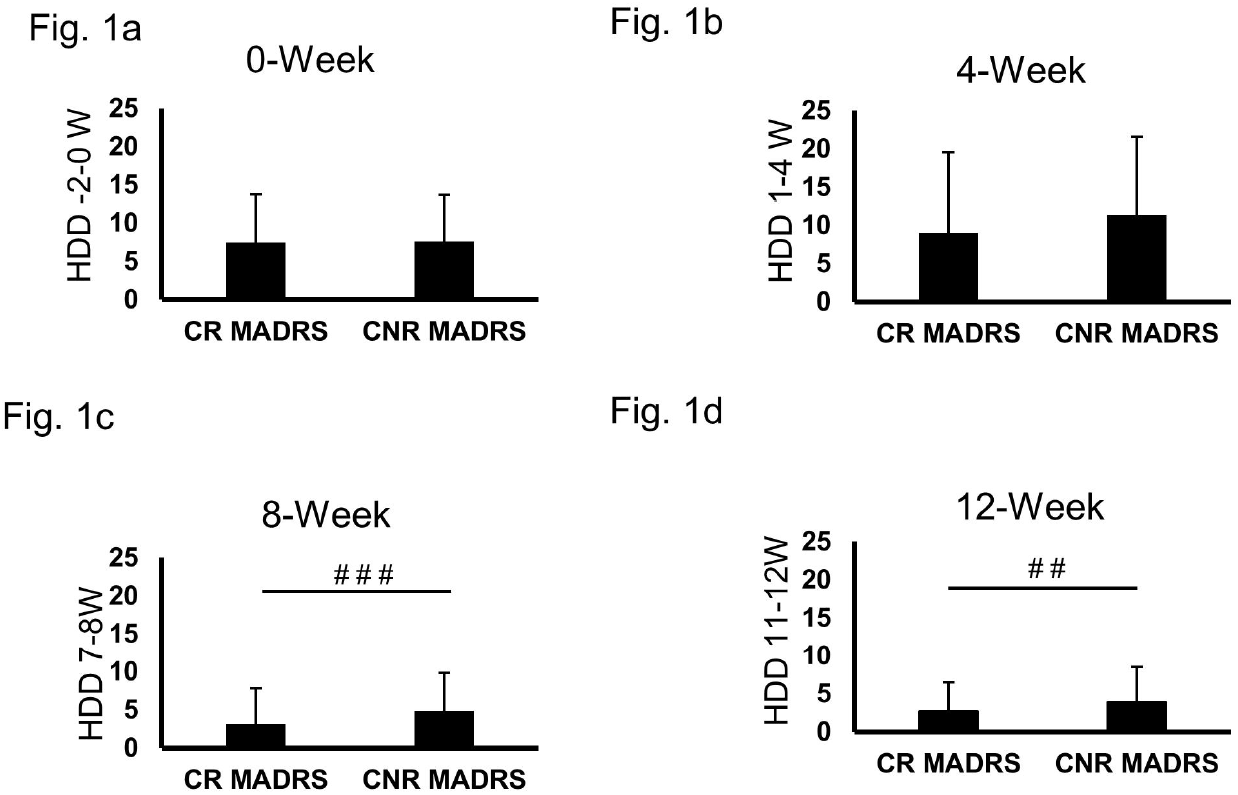
Changes in heavy drinking marker, heavy drinking days (HDD) in alcohol dependent patients receiving quetiapine as treatment, grouped as with clinically relevant MADRS (CR) and without any (CNR) recorded at each of the following timepoint: week 0 (Fig. 1a), week 4 (Fig. 1b), week 8 (Fig. 1c), and week 12 (Fig. 1d). With covariate MADRS score ^###^p = 0.001; and ^##^p=0.007. Data presented as Mean±SD. Statistical significance was set at p≤0.05.

**Figure 2:**
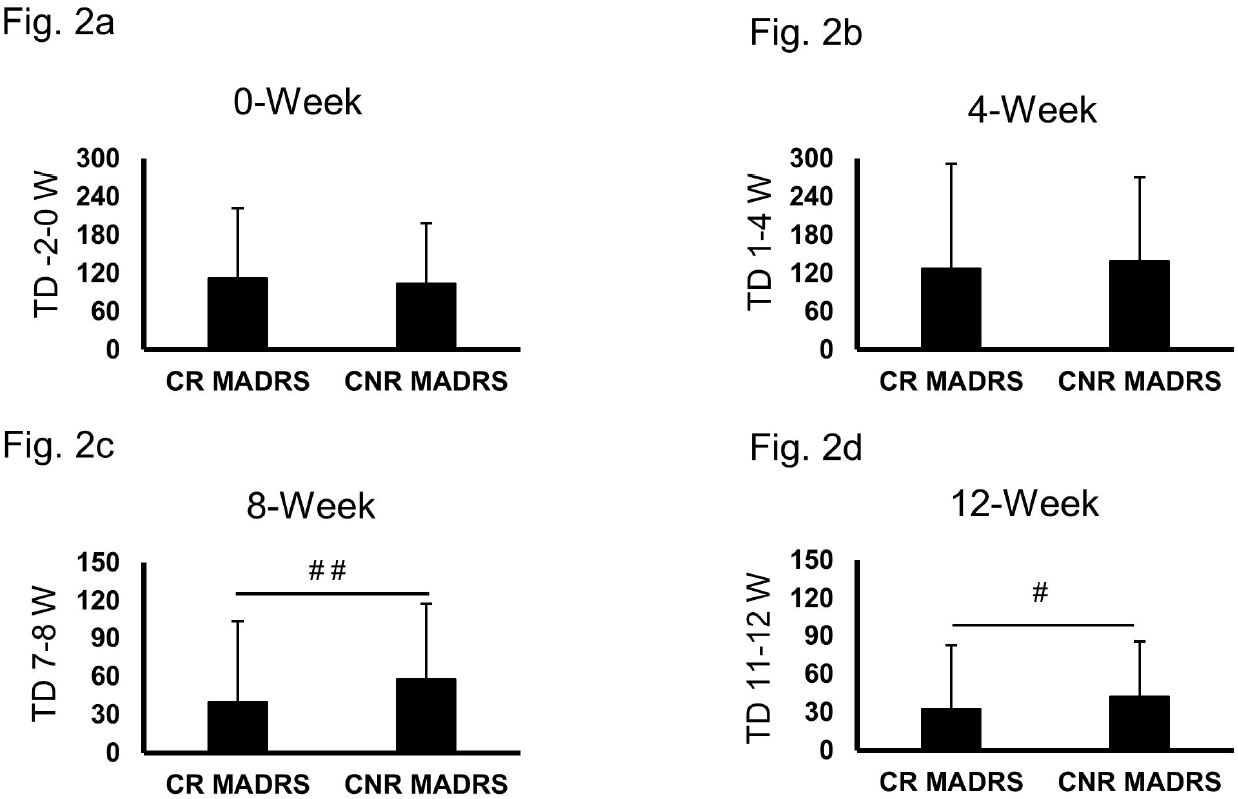
Changes in heavy drinking marker, Total drinks (TD) in alcohol dependent patients receiving quetiapine as treatment, grouped as with clinically relevant MADRS (CR) and without any (CNR) recorded at each of the following timepoint: week 0 (Fig. 2a), week 4 (Fig. 2b), week 8 (Fig. 2c), and week 12 (Fig. 2d). With covariate MADRS score; ^##^p=0.010, ^#^p=0.020. Data presented as Mean±SD. Statistical significance was set at p≤0.05.

**Figure 3:**
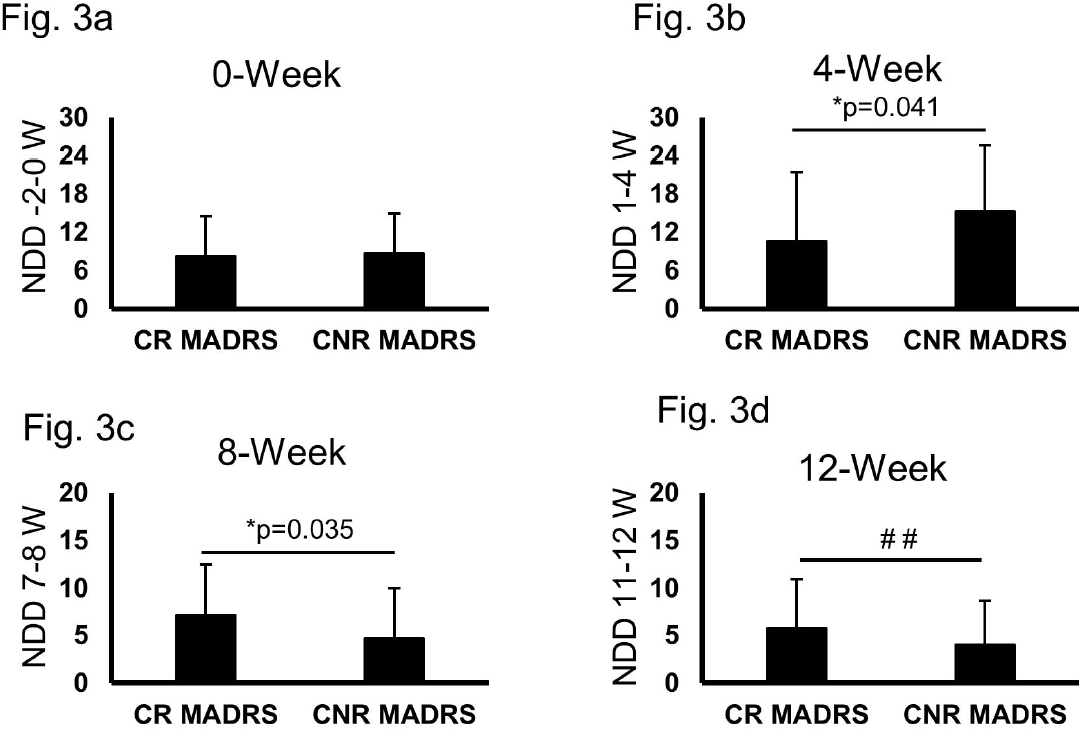
Changes in heavy drinking marker, number of days of drinking (NDD) in alcohol dependent patients receiving quetiapine as treatment, grouped as with clinically relevant MADRS (CR) and without any (CNR) recorded at each of the following timepoint: week 0 (Fig. 3a), week 4 (Fig. 3b), week 8 (Fig. 3c), and week 12 (Fig. 3d). With covariate MADRS score; and ^##^p=0.006. Data presented as Mean±SD. Statistical significance was set at p≤0.05.

**Figure 4:**
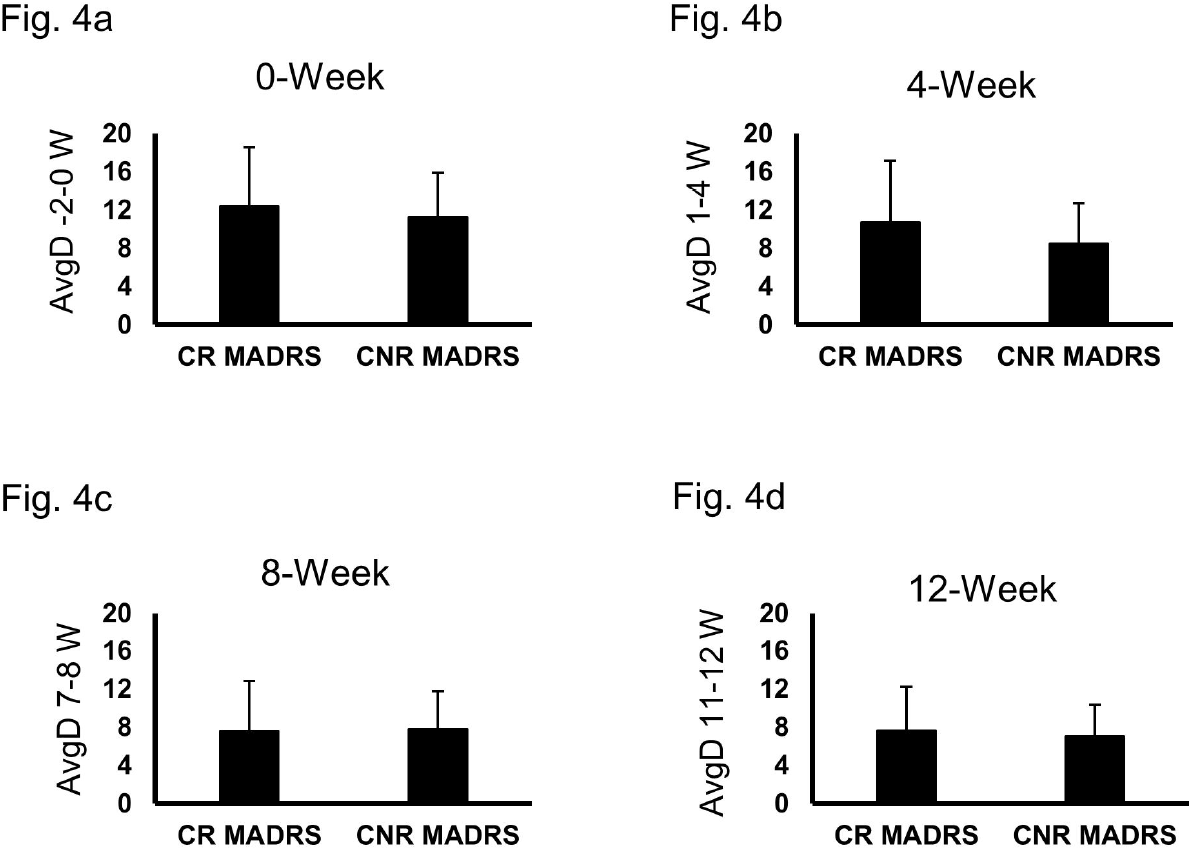
Changes in heavy drinking marker, average drinks (AvgD) in alcohol dependent patients receiving quetiapine as treatment, grouped as with clinically relevant MADRS (CR) and without (CNR) recorded at each of the following timepoint: week 0 (Fig. 4a), week 4 (Fig. 4b), week 8 (Fig. 4c), and week 12 (Fig. 4d). Data presented as Mean±SD. Statistical significance was set at p≤0.05.

HDD (7.45±6.2 at baseline vs. 2.68± 3.8 at 12 W) was lower in CR group compared to the CNR group (7.61±6.1 at baseline vs. 3.87± 4.6 at 12 W) (Fig. 1a and 1d). On average, this drop was around 1.34 heavy drinking episode per week (∼HDD) at 12 week. We found a significant within-subjects contrast (p≤0.001) in the HDD values across all the four timelines between CR and CNR groups. HDD values gradually decreased numerically in the CR group with highest changes observed at 8 week (3.1±4.7) vs. CNR (4.87±5.0) (Fig. 1c). HDD values were significantly lower at 8W (Fig. 1c) and 12W (Fig. 1d) in the CR group vs. CNR group when co-varied with the MADRS scores.

Progressive lowering of TD by time and treatment was well observed in the CR group (111.89±109.6 at baseline vs. 32.72±43.2 at 12 W) compared to the CNR group (103.53±94.9 at baseline vs. 42.36±49.9 at 12 W) (Fig. 2, all subfigures). On average, this drop was around 16.36 drinks per week (∼TD) at 12 week timepoint in the CR group. A significant within-subjects contrast was observed also in the TD values (p≤0.001) across all the four time points between CR and CNR groups when we used repeated measures ANOVA analysis. TD values were significantly lower at 8W (Fig. 2c) and 12 W (Fig. 2d) in the CR group vs. CNR group with MADRS scores being co-varied.

Baseline NDD (8.26±6.3) lowered initially at 4 week (end of drug titration, Fig. 3b) then increased at 8 week (Fig. 3c). It eventually decreased to 5.75±5.2 at 12 week (Fig. 3d) in the CR group. NDD showed similar response in the CNR group by time and treatment (6.3±6.1 at baseline vs. 3.8±4.6 at 12 W) (Fig. 3). CR group patients drank more frequently but in moderation (reduced heavy drinking days [HDD] and total drinks [TD] as mentioned before). There was a significant interaction effect of NDD values across all the four timepoints by both the groups, p=0.026 (along with a main effect: p<0.001 across all the four time points). Higher NDD levels were statistically significant at 8 W (p=0.035 [when co-varied with MADRS augmented p=0.004]) (Fig. 3c); and 12 W (non-significant p=0.106 [when co-varied with MADRS augmented p = 0.006]) (Fig. 3d) compared to the CNR group.

AvgD (12.38±6.2 at baseline vs. 7.65±4.6 at 12 week) remained higher in the CR group compared to the CNR group (11.25±4.6 at baseline vs. 7.06±3.3 at 12 week) by the end of the treatment (Fig. 4a and 4d). Average drinking remained higher in the CR group at week 4 of the treatment due to lower number of drinking days, when total drinks were not significantly different between the two groups (Fig. 4b). At 8-week, number of drinking days went up coupled with lowering of total drinks (Fig. 2c and Fig. 3c), which likely brought the average drinks to slightly lower level in CR group, this trend persisted till the end of treatment (Fig. 4c).There was a significant interaction effect of time in the average drinks per drinking day (AvgD) values between the CR and CNR groups, p=0.038; and a main effect of AvgD values, p<0.001 across all the four times. AvgD changes with treatment showed a numerical lowering in CNR group at each time (Fig. 4a-d), similar to NDD values. However, none of the comparisons between the CR and CNR groups for the AvgD marker were statistically different.

We assessed both numerically and statistically the values of the drinking markers in CNR and CR groups between the treatment and placebo arms for the last two week (11^th^-12^th^) of treatment (Table 2 and Fig. 5). We found lowering of TD, HDD and NDD in the CR group of quetiapine treated patients, whereas same markers showed higher numerical values in the CR group of placebo treated patients. There was not much variability found in AvgD values across the CNR and CR groups of each arm. In placebo arm, TD value remained at more than 20 drinks per week in the CR group at the end of the treatment assessment (Table 2). We verified this lowering by using AUC-ROC response (Fig. 5). We found that the response was positive consistent with the lowering (Table 2), however it was weak only (0.607) at the study end compared to baseline (0.518) in CR placebo group with respect to the CR quetiapine group patients.

**Table 2:** Drinking patterns reported in treatment and placebo arms in NCR and CR (MADRS) groups.

| Drinking Markers | Quetiapine arm |  | Placebo arm |  |
| --- | --- | --- | --- | --- |
|  | CNR group | CR group | CNR group | CR group |
| **TD 11-12W** | 42.35 ± 49.94 | 32.73 ± 43.17 | 33.49 ± 45.63 | 42.19 ± 50.23 |
| **AvgD 11-12W** | 7.06 ± 3.29 | 7.64 ± 4.63 | 6.85 ± 4.23 | 7.43 ± 3.86 |
| **HDD 11-12W** | 3.87 ± 4.62 | 2.68 ± 4.62 | 2.72 ± 3.86 | 3.52 ± 4.48 |
| **NDD 11-12W** | 5.75 ± 5.17 | 4.00 ± 4.65 | 4.63 ± 5.20 | 5.50 ± 5.20 |

TD 11-12W: Total drinks in 11 - 12 weeks; AvgD 11-12W: Average drinks per drinking day in 11 – 12 weeks; HDD 11-12W: heavy drinking days in 11 - 12 weeks; NDD90 11-12W: number of drinking days in 11 – 12 weeks.

**Figure 5:**
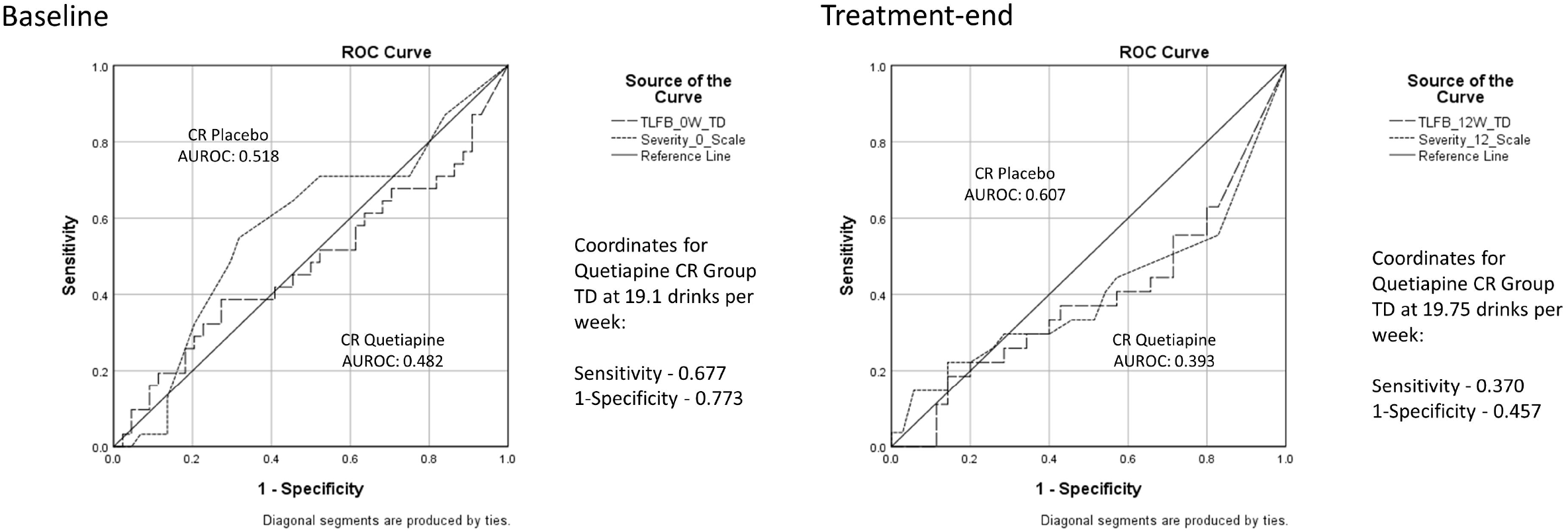
Drinking profile assessment for total drinks at baseline and treatment- end for the sub-groups categorized by the baseline reported MADRS (treated with Quetiapine or placebo) using area under the curve plot. Coordinates for TD (at ∼ 20 drinks per week) has also been mentioned for both the timelines.

### Association of heavy drinking and depression in Quetiapine treated AD Patients

There was a significant association (adjusted R^2^=0.162 at p=0.024) of depression ratings and heavy drinking days (HDD) at 4 weeks assessment in CR patients. This association grew progressively stronger over the treatment course, with the effect size increasing to adjusted R^2^ = 0.801 at p≤0.001 at week 12 (Table 3). In addition, over the same period in the CR group, significant associations with increasing effect sizes were observed for all other measures of drinking: total drinks, number of drinking days, and average drinks per day (Table 3). Increasing effect sizes were significantly observed with TD (adjusted R^2^=0.403 at 8 week to adjusted R^2^=0.776 at 12 week), and NDD (adjusted R^2^=0.294 at 8 week to adjusted R^2^=0.497 at 12 week). Such increasing effect sizes in patients with high MADRS support the fact that the efficacy of active treatment in heavy drinkers goes along with the greater association of both the lowering of drinking markers and the lowering of depressions scores. No such association was observed in CNR group patients (Table 3).

**Table 3:**
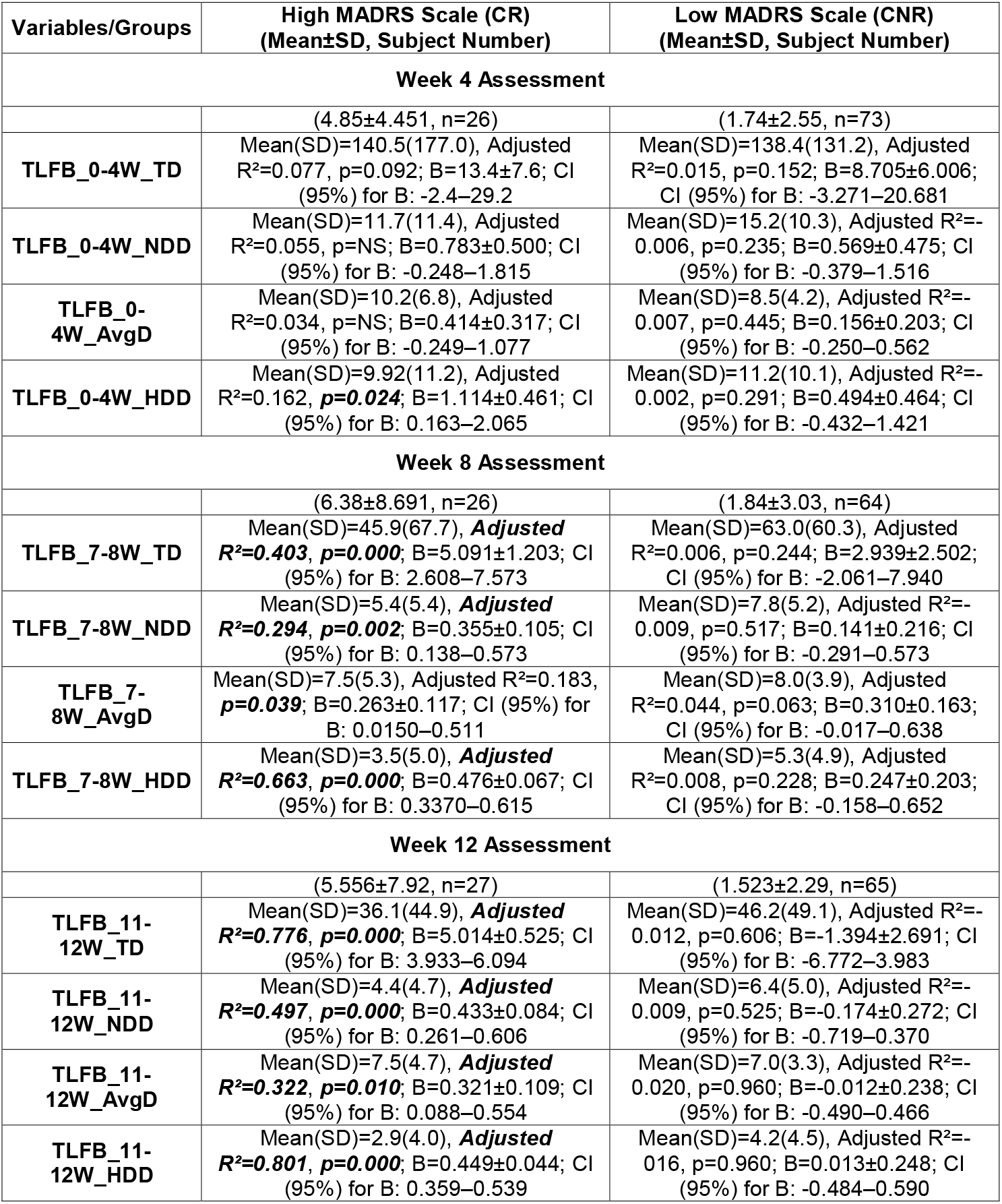
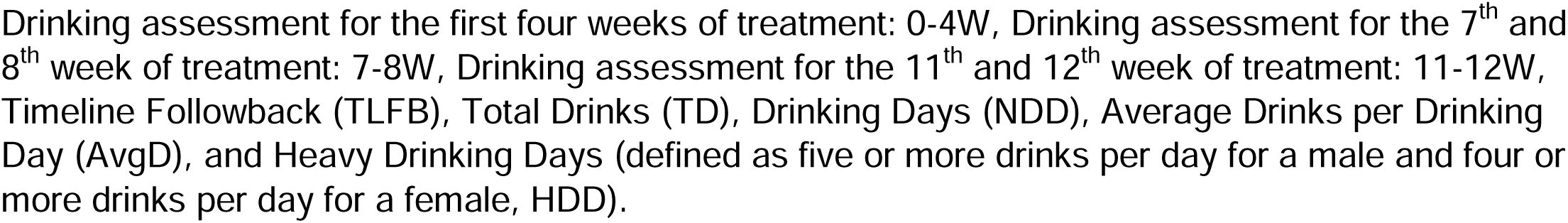
Regression analysis of drinking markers at each timepoint and baseline CR and CNR MADRS scores in patients treated with Quetiapine XR.

| Variables/Groups | High MADRS Scale (CR)<br>(Mean±SD, Subject Number) | Low MADRS Scale (CNR)<br>(Mean±SD, Subject Number) |
| --- | --- | --- |
| **Week 4 Assessment** |  |  |
|  | (4.85±4.451, n=26) | (1.74±2.55, n=73) |
| TLFB_0-4W_TD | Mean(SD)=140.5(177.0), Adjusted R<sup>2</sup>=0.077, p=0.092; B=13.4±7.6; CI (95%) for B: -2.4–29.2 | Mean(SD)=138.4(131.2), Adjusted R<sup>2</sup>=0.015, p=0.152; B=8.705±6.006; CI (95%) for B: -3.271–20.681 |
| TLFB_0-4W_NDD | Mean(SD)=11.7(11.4), Adjusted R<sup>2</sup>=0.055, p=NS; B=0.783±0.500; CI (95%) for B: -0.248–1.815 | Mean(SD)=15.2(10.3), Adjusted R<sup>2</sup>=-0.006, p=0.235; B=0.569±0.475; CI (95%) for B: -0.379–1.516 |
| TLFB_0-4W_AvgD | Mean(SD)=10.2(6.8), Adjusted R<sup>2</sup>=0.034, p=NS; B=0.414±0.317; CI (95%) for B: -0.249–1.077 | Mean(SD)=8.5(4.2), Adjusted R<sup>2</sup>=-0.007, p=0.445; B=0.156±0.203; CI (95%) for B: -0.250–0.562 |
| TLFB_0-4W_HDD | Mean(SD)=9.92(11.2), Adjusted R<sup>2</sup>=0.162, **p=0.024**; B=1.114±0.461; CI (95%) for B: 0.163–2.065 | Mean(SD)=11.2(10.1), Adjusted R<sup>2</sup>=-0.002, p=0.291; B=0.494±0.464; CI (95%) for B: -0.432–1.421 |
| **Week 8 Assessment** |  |  |
|  | (6.38±8.691, n=26) | (1.84±3.03, n=64) |
| TLFB_7-8W_TD | Mean(SD)=45.9(67.7), **Adjusted R<sup>2</sup>=0.403, p=0.000**; B=5.091±1.203; CI (95%) for B: 2.608–7.573 | Mean(SD)=63.0(60.3), Adjusted R<sup>2</sup>=0.006, p=0.244; B=2.939±2.502; CI (95%) for B: -2.061–7.940 |
| TLFB_7-8W_NDD | Mean(SD)=5.4(5.4), **Adjusted R<sup>2</sup>=0.294, p=0.002**; B=0.355±0.105; CI (95%) for B: 0.138–0.573 | Mean(SD)=7.8(5.2), Adjusted R<sup>2</sup>=-0.009, p=0.517; B=0.141±0.216; CI (95%) for B: -0.291–0.573 |
| TLFB_7-8W_AvgD | Mean(SD)=7.5(5.3), Adjusted R<sup>2</sup>=0.183, **p=0.039**; B=0.263±0.117; CI (95%) for B: 0.0150–0.511 | Mean(SD)=8.0(3.9), Adjusted R<sup>2</sup>=0.044, p=0.063; B=0.310±0.163; CI (95%) for B: -0.017–0.638 |
| TLFB_7-8W_HDD | Mean(SD)=3.5(5.0), **Adjusted R<sup>2</sup>=0.663, p=0.000**; B=0.476±0.067; CI (95%) for B: 0.3370–0.615 | Mean(SD)=5.3(4.9), Adjusted R<sup>2</sup>=0.008, p=0.228; B=0.247±0.203; CI (95%) for B: -0.158–0.652 |
| **Week 12 Assessment** |  |  |
|  | (5.556±7.92, n=27) | (1.523±2.29, n=65) |
| TLFB_11-12W_TD | Mean(SD)=36.1(44.9), **Adjusted R<sup>2</sup>=0.776, p=0.000**; B=5.014±0.525; CI (95%) for B: 3.933–6.094 | Mean(SD)=46.2(49.1), Adjusted R<sup>2</sup>=-0.012, p=0.606; B=-1.394±2.691; CI (95%) for B: -6.772–3.983 |
| TLFB_11-12W_NDD | Mean(SD)=4.4(4.7), **Adjusted R<sup>2</sup>=0.497, p=0.000**; B=0.433±0.084; CI (95%) for B: 0.261–0.606 | Mean(SD)=6.4(5.0), Adjusted R<sup>2</sup>=-0.009, p=0.525; B=-0.174±0.272; CI (95%) for B: -0.719–0.370 |
| TLFB_11-12W_AvgD | Mean(SD)=7.5(4.7), **Adjusted R<sup>2</sup>=0.322, p=0.010**; B=0.321±0.109; CI (95%) for B: 0.088–0.554 | Mean(SD)=7.0(3.3), Adjusted R<sup>2</sup>=-0.020, p=0.960; B=-0.012±0.238; CI (95%) for B: -0.490–0.466 |
| TLFB_11-12W_HDD | Mean(SD)=2.9(4.0), **Adjusted R<sup>2</sup>=0.801, p=0.000**; B=0.449±0.044; CI (95%) for B: 0.359–0.539 | Mean(SD)=4.2(4.5), Adjusted R<sup>2</sup>=-0.016, p=0.960; B=0.013±0.248; CI (95%) for B: -0.484–0.590 |

Drinking assessment for the first four weeks of treatment: 0-4W, Drinking assessment for the 7<sup>th</sup> and 8<sup>th</sup> week of treatment: 7-8W, Drinking assessment for the 11<sup>th</sup> and 12<sup>th</sup> week of treatment: 11-12W, Timeline Followback (TLFB), Total Drinks (TD), Drinking Days (NDD), Average Drinks per Drinking Day (AvgD), and Heavy Drinking Days (defined as five or more drinks per day for a male and four or more drinks per day for a female, HDD).

Quetiapine has sedative effects, thus we also tested if the results are not due to quetiapine acting primarily on sleep itself (by significantly augmenting the MADRS scores). In CR patients (with clinically relevant baseline MADRS scores), the raw scores reported for sleep score (1 item in the MADRS scale) was significantly associated with heavy drinking markers only at 8 week assessment (albeit effects were mild or low): HDD (adjusted R^2^=0.259, p=0.005), TD (adjusted R^2^=0.252, p=0.005), and NDD (adjusted R^2^=0.179, p=0.018). However, there was no association of sleep score and heavy drinking measures at baseline, 4-week and specially at the end of the study (12 week). Similarly, we did not find any significant association of heavy drinking measures and depression scale at each timepoint in CNR group.

## Discussion

At the baseline, 31 out of 108 alcohol dependent patients showed clinically relevant depression based on the MADRS scores in the active treatment group, and 44 in the placebo arm. None of the drinking markers showed any obvious differences of baseline drinking between the two groups nor were there differences in demographics. Furthermore, patients in our study who were prescribed antidepressants at least 30 days prior to the study were also eligible, who might show some elevation during the treatment/evaluation course. Even though the patients in this study were not diagnosed with depression based on the DSM-IV TR criteria, they exhibited some symptoms of depression, which is anticipated in heavy drinkers. ^26^ Thus, MADRS which does not diagnose depression, but measures intensity of certain symptoms was employed.

Clozapine has been reported to reduce alcohol use in schizophrenic patients with alcohol use disorder. ^34,35^ In our treatment study, one group of alcohol dependent patients also exhibited clinically relevant symptoms of depression indicating presence of comorbid condition (CR). We found separation in the treatment efficacy of quetiapine based on the level of depression symptoms recorded at baseline. Both total drinks (by one-fourth) and heavy drinking days (by one third) continued to be reduced significantly over the treatment course. Average drinking and number of drinking days also dropped in the CR group compared to baseline, but levels were higher than those in the CNR group.

Quetiapine (Seroquel®), like clozapine, is another atypical antipsychotic medication that, in addition to decreasing alcohol intake, has also been reported to decrease craving and psychiatric symptoms in alcoholic patients with a concurrent axis I disorder ^36^ in a 4-month open label trial. In our findings, lowering of heavy drinking markers showed increasing effect sizes and significance of association when regressed with MADRS during the course of treatment only in CR patients. We also found a statistically and numerically significant elevation in total amount of drinking and the pattern of heavy drinking persisted at each time point until the end of the treatment in CNR patients. This response was in contrast to that observed in the CR group. Quetiapine might be an effective treatment in AD patients who also have depression symptoms (CR) but may not be effective in AD patients without any significant level of depression symptoms (CNR). Kampman and colleagues also reported a higher therapeutic value of quetiapine in treatment of Type B alcoholics, the more complex of the two types of alcoholism characterized by earlier onset of problematic drinking, more severe alcohol dependence, and greater psychopathology. ^25^

One study demonstrated that aripiprazole decreased drinking in alcoholic patients who were not seeking treatment ^37^ while another study showed that aripiprazole proved to be as effective as the currently FDA approved drug, naltrexone, in treating alcohol dependence. ^38^ A parent study of this manuscript reported that quetiapine could overall lower the drinking in AD patients on quetiapine, although not to a level that is considered to be moderate drinking. ^27^ Based on our findings, we observed that quetiapine selectively targets and lowers the alcohol consumption in alcohol dependents who also show baseline signs of depression. This suggests that drinking markers are good therapeutic targets to assess treatment efficacy and corresponding lowering of depression scores in heavy drinkers could be a positive sign of drug efficacy. Similar findings were also reported in a study comparing aripriprazole and placebo in a multicenter drug trial. ^39^ Scope of that study did not include investigating the results in context of the presence of depression symptoms. HDD at baseline was roughly four times per week that reduced to roughly one episode per week at the end of the treatment. Similarly, baseline NDD was four times a week that reduced to roughly three times per week. Importantly TD reduced from roughly 56 drinks per week at baseline to 16.5 drinks per week, which was lower than the heavy drinking criteria of 20 drinks per week for men (we had almost all male participants in the CR group [27/31]). ^18^ Thus, treatment efficacy of anti-depressants should also be evaluated in context of depression symptoms apart from overall outcomes. Efficacy of Quetiapine in lowering the heavy drinking patterns was evident in the subset of patients who exhibited baseline clinically relevant MADRS scores.

A clinical study showed that olanzapine reduced the urge to drink in heavy social drinkers post exposure to alcohol cues ^40^ as well as diminished alcohol craving and consumption in alcohol-dependent patients, especially in those with the 7-repeat allele of the D4 receptor gene. ^41^ In our study, we found a corresponding lowering in the number of drinking days during treatment) in CNR patients. While quetiapine could not lower drinking from heavy to moderate levels in this group, there was a non-significant lowering in total drinks at the 12-week assessment compared to baseline. Another study showed similar findings with no differences in drinking outcomes between olanzapine and placebo groups in alcohol dependent patients. ^42^ In that study, the authors used Beck’s Depression Inventory (BDI), however association between BDI and heavy drinking was not discussed. It seems that the potential effectiveness of an anti-depressant drug should be evaluated in the context of both lowering alcohol consumption and improving depression symptoms.

This study had several limitations. There were 3-fold more men in this study compared to women, which shifted balance of the sex ratio disproportionally. Though since this occurred in all the groups evenly, it did not restrain the analysis. There were a few dropouts during the course of the study and 4–5% of the total data points could not be included in this study due to missing values. In our study, AD patients with clinically relevant depression symptoms (−CR group) were far fewer (<50%) than the group who did not have clinically relevant depression symptoms (CNR group). This could have tilted the weight of findings toward the CNR group in the analyses. AUROC curve supports the positive response thus, we believe that the findings are still significant. There is a likelihood of type I error (false positive) results in multiple comparisons for TD and HDD comparisons. We see numerical drops regardless that were lower than the heavy drinking criteria, and ROC curve showed positive response for true positivity (This response was weak by category). We did not include MDD patients with comorbid diagnosis of alcohol in this study, which might have also revealed if Quetiapine has dual efficacy, however this was not part or scope of this study and its design. We did not evaluate sleep comprehensively (there is only one on sleep in the MADRS scale, which was not significantly associated in CR group apart from 8-week stage) for its role in the treatment ^30^. Scope of this study was limited to interaction of depression and drinking on treatment efficacy. In a separate study, we would address using a comprehensive questionnaire on the interaction of sleep and quetiapine in alcohol consumption (Pittsburg sleep quality index).

## Conclusions

Quetiapine fumarate XR reduced markers of heavy drinking, namely, (1) heavy drinking days and (2) total drinks at the end of treatment compared to baseline albeit only in alcohol dependent patients who exhibited clinically relevant depression symptoms at baseline (sub-set efficacy is evident that generalized efficacy). In AD patients without any relevant level of baseline signs of depression, quetiapine could reduce the frequency of drinking days during the treatment, however, little effect was observed in heavy drinking days and total drinks. Quetiapine fumarate treatment showed therapeutic dimorphism in alcohol dependent patients and could be an effective intervention for those who have clinical signs of depression along with heavy alcohol intake.

## Data Availability

The datasets during and/or analyzed during the current study available from the corresponding author on reasonable request.

## Declarations

### Authors’ contribution

VV is the project PI and designed the study. VV, SK, MLS and LMM provided data acquisition, management and analysis. VV, KVC, VAR, NS and CJM interpreted the outcomes. VV, KVC, LMM and CJM wrote the manuscript. VV, VAR, KVC, CJM, NS, SK, and MLS contributed scientifically. All the authors have approved the submission version of this manuscript.

### Proprietorship

This article is a work of the University of Louisville Alcohol Research Center and is in the public domain in the USA.

## Acknowledgments

We thank research/clinical staff of NIAAA; and research staff of the University of Louisville for their support. We thank Marion McClain for editorial support for this manuscript.

## Ethics approval and consent to participate

Study was approved by the site/s’ Institutional Review Board (Ethics committee). All patients included in this study consented to participate before the beginning of the study.

## Abbreviations

AD: Alcohol dependents
AUD: Alcohol use disorder
CR: Clinically relevant
CNR: Clinically not-relevant
MADRS: Montgomery-Asberg Depression Rating Scale
XR: Extended Release
TLFB90: Timeline Follow back past 90 days
TD90: Total drinks in 90 days
AvgD90: Average drinks in last 90 days
HDD90: Heavy drinking days in last 90 days
NDD90: Number of drinking days in last 90 days
TD: Total Drinks during specific treatment-interval
NDD: Drinking Days during specific treatment-interval
AvgD: Average Drinks per Drinking Day during specific treatment-interval
HDD: Heavy Drinking Days during specific treatment-interval.

